# Relative disease burdens of COVID-19 and seasonal influenza in New York City, February 1 - April 18, 2020

**DOI:** 10.1101/2020.04.22.20073551

**Authors:** Jeremy Samuel Faust, Carlos del Rio

## Abstract

Comparisons between the mortality burdens of COVID-19 and seasonal influenza often fail to account for the fact that the United States Centers for Disease Control and Prevention (CDC) reports annual influenza mortality estimates which are calculated based upon a series of assumptions about the underreporting of flu deaths. COVID-19 deaths, in contrast, are being reported as raw counts. In this report, we compare COVID-19 death counts to seasonal influenza death counts in New York City during the interval from February 1 - April 18, 2020. Using this approach, COVID-19 appears to have caused 21.4 times the number of deaths as seasonal influenza during the same period. We also assessed excess mortality in order to verify this finding. New York City has had approximately 13,032 excess all-cause mortality deaths during this time period. We assume that most of these deaths are COVID-19 related. We therefore calculated the ratio of excess deaths (i.e. assumed COVID-19 deaths) to seasonal influenza deaths during the same time interval and found a similar ratio of 21.1 COVID-19 to seasonal influenza deaths. Our findings are consistent with conditions on the ground today. Comparing COVID-19 deaths with CDC estimates of yearly influenza-related deaths would suggest that, this year, seasonal influenza has killed approximately the same number of Americans as COVID-19 has. This does not comport with the realities of the pandemic we see today.

## Introduction

Some experts now predict that COVID-19 will cause approximately 60,000 deaths in the United States by August of 2020, seemingly similar to seasonal influenza mortality statistics in recent years. However, such comparisons fail to account for differences in how deaths from SARS-CoV-2 / COVID-19 and seasonal influenza are reported. Seasonal influenza mortality is reported by the Centers for Disease Control and Prevention (CDC) as adjusted estimates, and also includes pneumonia deaths caused by other pathogens. Between 2010 and 2019, the annual estimates of influenza-associated and pneumonia deaths ranged from 12,000 and 61,000 deaths per year [1, 2]. COVID-19 mortality, by contrast, is being tallied and reported as raw counts, and has caused 13,240 in New York City alone, through April 18, 2020 [3, 4].

A more valid comparison between SARS-CoV-2 / COVID-19 and seasonal influenza-associated mortality is possible by juxtaposing data obtained using similar methodologies--i.e. comparing raw death counts for both diseases. However, this may not convince even some, as the determination of a proximate cause of death that appears on death certificates rely upon physician judgment and is prone to error [5]. Therefore, the New York City novel coronavirus outbreak provides an unusual opportunity. According to the CDC there were approximately 13,032 excess deaths in New York City between February 1 and April 18, as compared to the same interval in 2017-2019. It is reasonable to assume that these deaths were attributable to COVID-19. Here, we compare the excess deaths in New York City to seasonal influenza counted deaths over the 76-day interval, as well as counted COVID-19 deaths as reported by the CDC and the New York Department of Health and Mental Hygiene (NYDOH).

## Methods

Mortality statistics were obtained from the CDC and the National Center for Health Statistics (NCHS) and the NYDOC. Counted influenza deaths (ICD J09-J11) as reported to and by the CDC/NCHS during the interval from February 1 to April 18, 2020 were compared to: 1) the CDC‚s reported number of excess deaths in New York city; 2) the CDC‚s reported counted number of deaths due either to confirmed or suspected COVID-19 (ICD UO7.1) and; 3) the NYDOH reported counted number of confirmed and probable COVID-19 deaths. Mortality ratios were calculated.

## Results

The CDC reports that between February 1 and April 18, the number of all-cause deaths was 171% of expected usual figures compared to the same interval in 2017-2019. The number of New York City confirmed and “total” COVID-19 deaths (confirmed COVID-19 deaths plus probable COVID-19 deaths, as reported by the NYDOH) was 21.4 times greater than the number of counted influenza deaths reported by the CDC during the same interval (Table 1). The ratio of all-cause excess mortality in New York City to influenza deaths during this time was 21.1. The ratio of counted COVID-19 deaths (based on ICD codes reported to and by the CDC) was 9.5 times greater than influenza over the same time period.

**Table 1.** New York City Deaths, February 1 - April 18, 2020.

| Source | Deaths |
| --- | --- |
| CDC reported influenza counted deaths. | 619* |
| CDC reported excess deaths, all-cause mortality. | 13,032* |
| CDC reported COVID-19 counted deaths (ICD U07.1). | 5,870* |
| NYDOH reported COVID-19 total deaths. | 13,240 ^ |
| NYDOH reported COVID-19 confirmed deaths. | 8,811 ^ |
| NYDOH reported COVID-19 probable deaths. | 4,429 ^ |

\*CDC, https://www.cdc.gov/nchs/nvss/vsrr/COVID19/index.htm.
^NYDOH, https://www1.nyc.gov/assets/doh/downloads/pdf/imm/covid-19-deaths-confirmed-probable-daily-04192020.pdf (first death, March 11).

## Discussion

Public officials and health and policy experts frequently quote CDC statistics that indicate that nearly 61,000 Americans per year have died of influenza during the last seven seasons. However, these are calculated estimates based upon numerous assumptions, all of which reinforce the belief that influenza deaths are undercounted at multiple points in the healthcare system, from under testing, to underreporting of hospitalization, to under attribution on death certificates.

During the current COVID-19 outbreak, approximately 40,000 Americans have died and thousands more are expected to die in the coming months. Meanwhile, the CDC states that this year, between 24,000 and 62,000 Americans have died of influenza [7]. Placed side-by-side, the absurdity of these numbers is self-evident; conditions on the ground do not support statistics that suggest that seasonal influenza has killed approximately the same if not many more Americans than COVID-19 has. Instead, by abandoning the statistical misadventure of estimating influenza deaths, and instead simply relying on reported counts, a far better quantitative and qualitative portrait of the relative mortality burdens of SARS-CoV-2 / COVID-19 and seasonal influenza emerges.

This study has several limitations. First, while counted deaths are likely to be more accurate than estimated deaths, both influenza and COVID-19 deaths may either be under or overreported. In the case of COVID-19 deaths, it remains unclear who died *from* SARS-CoV-2 infection and who died *with* the infection. However, the same forces can be applied to influenza counted deaths. Second, while far lower than calculated estimates, influenza counted deaths may yet still be overestimations, as ICD codes used (J09-J11) actually permit and do not exclude COVID-19 and pneumonia (caused by other pathogens) from appearing on death certificates in which influenza was determined to be the cause of death [3]. Third, while the 13,032 excess deaths in New York City since February 1st may reasonably be attributed to COVID-19, this supposition cannot at present be explicitly proven. Equally, however, it may be that the number of COVID-related deaths is in fact greater than the number of excess deaths, as deaths from other causes, such as trauma and even heart attacks, appears to have decreased during the shelter-in-place period of the pandemic [6]. Fourth, the lower ratio of counted COVID-19 to influenza deaths calculated using CDC numbers (in lieu of NYDOH figures) likely reflects lags in COVID-19 related deaths, delays which the agency acknowledges openly. Such lags are less likely to affect influenza deaths, as the peak of the season appears to have occurred in February. Finally, influenza deaths have occurred throughout the 76-day period in this study. The first COVID-19 death in New York City did not occur until March 11. Therefore, the ratios we report here may in fact underestimate the relative mortality burden of COVID-19. The CDC has stated that week-to-week state-level data for 2020 may soon be published, which would allow comparisons of the relative burdens of these two diseases at their peaks.

In conclusion, comparing actual influenza and COVID-19 deaths in the same city suggests that COVID-19 mortality burden is indeed an order of magnitude greater than that of seasonal influenza.

## Data Availability

All data are available on the websites of the United States Centers for Disease Control and Prevention, and the New York City Department of Health and Mental Hygiene.

http://www.cdc.gov

https://www1.nyc.gov/site/doh/about/about-doh.page

